# Ecological Momentary Assessments of daily pain experiences in bothersome and high-impact chronic pain

**DOI:** 10.64898/2026.03.18.26348727

**Authors:** Marta Walentynowicz, Doerte U. Junghaenel, Sean C. Mackey, Michael Von Korff, Arthur A. Stone

**Author notes:** Corresponding Author:* Marta Walentynowicz, Dornsife Center for Self-Report Science and Center for Economic C Social Research, University of Southern California, 635 Downey Way suite 405, Los Angeles, CA 90089, USA. Presentation: Results of this study were presented at the 83rd Annual Meeting of the Society for Biopsychosocial Science and Medicine (Chicago, IL, USA, March 2026).

## Abstract

High-impact chronic pain (HICP), defined as persistent pain that substantially limits daily activities, affects millions of adults and poses a public health challenge. Yet relatively little is known about how HICP manifests in people’s daily lives. To address this gap, this study used the comprehensive Ecological Momentary Assessment of pain (cEMAp) to assess pain-related experiences four times per day over 7 days in individuals with chronic low back pain. Based on the classification using the Graded Chronic Pain Scale–Revised, we compared individuals with HICP (*n* = 66) with those in the next most severe pain category, bothersome chronic pain (*n* = 41), defined as having similar pain frequency but less frequent interference with daily activities. On each prompt, participants completed 2-hour assessments of pain intensity, interference, catastrophizing, behaviors, coping strategies, and pain characteristics. In line with prior research, both groups reported similar pain intensity levels, but the HICP group reported more frequent interference with physical, mental, and social activities. There were no group differences in daily mood or catastrophizing. Exploratory analyses suggested that many daily experiences were similar across groups, with differences observed in selected pain qualities, coping strategies, and pain behaviors. Additional analyses of response distributions showed some similarity across groups in many experiences. Overall, although individuals with HICP on average experience higher pain interference in daily life, levels of many day-to-day experiences are similar between the two groups. Data obtained with cEMAp complement traditional retrospective assessment by providing a detailed view of chronic pain in everyday life.

## Introduction

High-impact chronic pain (HICP), defined as persistent pain lasting 3 months or more with significant impact on daily life or work activities [33], is a major public health challenge affecting approximately 8.5% of U.S. adults [6,15]. Compared to less severe chronic pain, HICP is characterized by greater limitations in mobility, self-care, and increased psychological burden, often resulting in disability, employment loss, and reduced quality of life [10,21,32]. Because HICP represents the most burdensome end of the chronic pain spectrum, early identification and treatment are crucial to address its rising prevalence and prevent long-term individual and societal burden.

Despite growing interest, relatively little is known about how HICP manifests in people’s daily lives. This is noteworthy because HICP is defined by pain and activity limitations occurring “on most days or every day” [21], yet most evidence on HICP is from retrospective assessments that are prone to recall bias and memory heuristics [26,29,31]. These methods also require aggregation of experiences over extended periods, possibly distorting the measurement of everyday pain experiences [3,25,28,29], and obscuring clinically relevant short-term fluctuations in pain intensity and impact [22–24,27].

Repeated momentary assessments offer an alternative to retrospective self-reports by providing fine-grained, temporally dense pictures of individuals’ daily experiences. Ecological Momentary Assessment (EMA) [22,24,27,29], a data capture procedure wherein individuals rate their experiences repeatedly throughout the day, minimizes recall bias, enables detection of temporal patterns, and facilitates new ways of quantifying pain-relevant experiences [22,24,29]. EMA is, therefore, ideally suited to study the experiences of individuals with chronic pain in their natural environments.

The present study utilizes data from the comprehensive Ecological Momentary Assessment of pain (cEMAp), a novel tool using a biopsychosocial approach to characterize individuals’ chronic pain experience in daily life [12]. cEMAp captures pain and pain-relevant dimensions for the previous 2 hours multiple times daily over one or more weeks, enabling fine-grained tracking of pain variability, daily fluctuations, and contextual factors typically missed in retrospective assessments.

This study aims to shed light on the daily lives of individuals with chronic pain using high-resolution, repeated assessments collected over one week. Participants were classified into four chronic pain categories using the Graded Chronic Pain Scale–Revised [32]. The present paper focuses on the contrast between bothersome and HICP because of their distinct clinical significance and treatment urgency. These two groups are characterized by pain on most or every day but differ in the frequency of pain interference. Prior findings suggest that individuals with HICP show substantially greater pain interference and activity limitation, as well as elevated depression, anxiety, and pain catastrophizing compared to those with bothersome chronic pain [30,32]. We therefore expected average group differences in daily measures of these constructs. Given prior work, we did not expect differences in momentary pain intensity. Other domains, including pain descriptors, locations, coping strategies, and behaviors, were examined on an exploratory basis to present a full picture of individuals’ daily functioning. Sex and age were controlled for as potential confounders to more precisely characterize group differences in daily experiences [13,14].

## Method

### Participants

The data for this cEMAp study are from an NIH-funded R61 project (R61NS118651; S.M., P.I.) designed to discover and validate composite diagnostic and prognostic biomarker signatures of musculoskeletal HICP following usual multidisciplinary pain treatment. The R61 enrolled and characterized adult participants at baseline and at 1-, 2-, 3-, and 6-month follow-ups. For this study, only the baseline data were used. Recruitment was conducted through the Stanford Pain Management Center (SPMC), including clinician referral/clinic outreach and the Stanford learning health system (CHOIR) research contact pathways.

Participants were eligible if they were aged 18–80, had chronic musculoskeletal pain (defined as pain occurring on at least half of days for ≥6 months), were fluent in English, able to complete study procedures, and represented a broad range of pain impact (PROMIS Pain Interference T-score of 40–80). Key exclusion criteria included inflammatory rheumatic disease or a non-musculoskeletal primary pain diagnosis, significant cognitive impairment, contraindication to magnetic resonance imaging, medical or psychiatric instability that could interfere with participation, current medicolegal factors (e.g., an open disability claim), plans for surgery during the follow-up period, and pregnancy. Study procedures were approved by the Stanford Institutional Review Board (Protocol #60657).

### Study Procedure

At the initial lab visit, participants provided informed consent, completed several tasks outside the scope of this paper, and received a link to complete a battery of questionnaires assessing their health and well-being online. They were also provided with written instructions about the EMA study. Starting from the day after the lab visit, participants received four prompts per day for 7 consecutive days. Prompts were delivered via text message and contained a link to a survey administered through the REDCap electronic data capture system hosted at Stanford University [8,9]. Each survey took approximately 5 minutes to complete and could be started within 1 hour of the prompt. To ensure broad coverage across waking hours, prompts were scheduled using a stratified random sampling approach that distributed survey times across four predefined 2-hour windows (8:00–9:59 AM, 12:00–1:59 PM, 4:00–5:59 PM, and 8:00–9:59 PM). Participants received financial compensation for completed study activities; travel reimbursement and ride-share support were provided as needed to minimize participation burden.

### Measures

The GCPS-R was used to categorize participants by chronic pain impact [32]. The GCPS-R is a validated five-item instrument that distinguishes among *no, mild, bothersome,* and *high-impact* chronic pain. The first two items assess the frequency of pain and associated activity limitations during the past 3 months (*never, some days, most days,* or *every day*). Participants who report pain *never* or on *some days* are classified as no chronic pain. Individuals experiencing pain on *most days* or *every day* are considered to have chronic pain and are further classified into the subsequent three groups. Those who also report that pain limits daily or work activities on *most days* or *every day* are classified as having HICP. Among the remaining individuals with chronic pain, the total score on three 0– 10 items—average pain intensity, interference with enjoyment of life, and interference with general activities (the PEG score)—determines whether chronic pain is bothersome (PEG ≥ 12) or mild (PEG < 12). The GCPS-R also includes an additional yes/no item assessing ability to work due to pain, which is not part of the scoring algorithm.

### cEMAp

The cEMAp is a multidimensional electronic diary instrument grounded in the biopsychosocial framework, designed to capture the physical, emotional, and social aspects of pain experience in daily life [12]. The measure integrates and adapts validated items from established legacy instruments to provide an ecologically valid assessment of pain in naturalistic settings. In developing the cEMAp, particular attention was given to selecting an appropriate timeframe. EMA studies in pain research have used both momentary (e.g., just before the prompt) and coverage (e.g., since the last prompt) approaches [16]. Although the former can minimize recall bias, it captures only a brief snapshot of the day and may overlook fluctuations or pain flares that occur between prompts. To enhance temporal coverage and characterize short-term variability in pain, cEMAp adopted a 2-hour recall timeframe, consistent with the coverage EMA model [35]. The cEMAp therefore employs a hybrid design, combining a single momentary rating with 2-hour recall assessments within each survey.

Each cEMAp survey began with a rating of momentary pain intensity on a numerical rating scale (NRS) ranging from 0 = *no pain at all* to 10 = *the worst imaginable pain*. For the first survey of the day, a brief morning module assessed sleep duration and sleep quality from the previous night on a 4-point scale ranging from 1 = *very good* to 4 = *very bad*. The 2-hour recall section assessed contextual factors (activities, location, social contact, temperature, stressful events) using multiple-choice questions, as well as mood on a 5-point scale (1 = *poor* to 5 = *excellent*) and presence of pain in the past 2 hours (yes/no). If participants reported experiencing pain in the past 2 hours, additional questions were administered, all referring to the pain experienced during that 2-hour window. Specifically, participants rated average, worst, and least pain intensity (0–10 NRS), pain duration (in minutes), and pain fluctuations. They then responded to items covering six additional domains: pain interference (8 items), pain catastrophizing (5 items), pain behavior (12 items), pain coping strategies (11 items), pain quality (16 items), and pain locations (19 items). For each domain, participants selected all items that applied. “Other” options allowed for open-text responses. Multiple-choice items were coded as 1 (endorsed) and 0 (not endorsed). The cEMAp survey questions have been previously published [12].

### Statistical Analyses

#### Sample Characteristics

Participants were included in the current analyses if they completed the GCPS-R at baseline and responded to at least 50% of EMA prompts during the baseline week. Based on GCPS-R scoring, participants were classified into four pain impact categories. The present analyses focused on those classified as having bothersome chronic pain and HICP, as these groups reflect clinically actionable levels of pain impact. Descriptive statistics for all four GCPS-R categories are provided in the online supplement. Demographic characteristics were compared between the bothersome chronic pain and HICP groups using Welch’s *t*-tests for continuous variables and chi-square tests for categorical variables.

#### Group Comparisons

EMA compliance was calculated separately for momentary and morning prompts as the proportion of assigned prompts completed (momentary: 28 prompts, morning: 7 prompts). Single continuous outcomes (i.e., pain intensity, mood, sleep quality) were aggregated at the person level across the sampling week prior to analyses. Scores for items asked at every prompt (i.e., momentary pain intensity and mood) were averaged across all completed prompts. Sleep quality was averaged across completed morning prompts. At each prompt, participants indicated whether they had experienced pain in the past 2 hours; when endorsed, they were asked to rate its intensity. The proportion of prompts with pain in the past 2 hours was computed as the number of prompts in which pain was endorsed divided by the total number of completed prompts. Because the intensity rating was prompted conditionally, two person-level indices were derived to capture different aspects of pain experience. The conditional 2-hour pain intensity score was calculated by averaging intensity ratings across prompts in which pain was endorsed, reflecting the average intensity of pain when pain occurred. The non-conditional 2-hour pain intensity score was calculated by averaging intensity ratings across all completed prompts, incorporating both pain intensity and frequency across the sampling week. Group differences in continuous outcomes were examined using analyses of covariance (ANCOVA) controlling for age and sex.

Items from pain-related domains (pain interference, pain catastrophizing, pain coping strategy use, pain behavior, pain quality, and pain location), each rated as present or absent, were analyzed using a two-stage approach to examine both overall domain-level differences and item-specific group differences.

First, a composite score was derived for each domain reflecting the number of endorsed items per prompt (e.g., the number of interference items or coping strategies). Endorsed items were summed within each prompt to create a domain count, which was then averaged across prompts in which pain was reported in the past 2 hours over the sampling week, yielding a person-level mean domain score.

Second, individual items within each domain were analyzed separately. Person-level proportions were computed for every item (absent coded as 0; present as 1), reflecting the frequency with which each item was endorsed across the sampling week. Because these questions were presented only when pain was reported in the past 2 hours, proportions were calculated relative to the number of prompts with pain, thereby accounting for individual differences in compliance and pain presence.

Each cEMAp domain (i.e., the set of item-level proportions within that domain) was then examined using a multivariate analysis of covariance (MANCOVA) controlling for age and sex to test for overall multivariate group effects. When the omnibus multivariate test was significant at *α* = .05, follow-up ANCOVAs were conducted for individual items within the domain. *p*-values for these follow-up tests were adjusted for multiple comparisons using the Benjamini–Hochberg false discovery rate (FDR) correction [2]. Effect sizes are reported as partial eta-squared (η²p) and interpreted using Cohen’s guidelines (small ≈ 0.01, medium ≈ .06, large ≈ .14) [4].

The homogeneity of covariance matrices was assessed using the Box test. Because this assumption was frequently violated, we adopted two complementary strategies to ensure robust inference. First, multivariate test statistics are reported using Pillai’s Trace, which is more robust to violations of normality and homogeneity than alternative multivariate statistics [17,20]. Second, we conducted permutation-based sensitivity analyses. For omnibus tests, a permutational multivariate analysis of variance (PERMANOVA) was performed with 9,999 permutations using the adonis2 function from the *vegan* R package [1]. For follow-up ANCOVAs, permutation-based tests were implemented using the Freedman–Lane method with 9,999 permutations (*permuco* R package [7]). When permutation and parametric analyses yielded consistent inferences, only parametric results are reported; otherwise, both are presented for transparency.

#### Analysis of Distributional Overlaps

To complement group comparisons, we examined the extent to which the distributions of the bothersome and HICP groups overlapped. Overlap was quantified as the area of commonality between the groups’ kernel density estimates of the person-level distributions [19]. The resulting overlapping index η can be interpreted as a similarity measure ranging from 0 (no overlap) to 1 (complete overlap) and can be applied to non-normal distributions. The overlapping index was computed using the *overlapping* R package [18].

#### Content Analysis of “Other” Responses

Finally, we examined the content of the “Other” response option across all six cEMAp domains. First, group differences between the bothersome and HICP groups in person-level proportions of “Other” endorsements were examined using ANCOVAs controlling for age and sex. For domains showing group differences, free-text responses were independently reviewed by two raters to identify recurring themes not represented in the existing response options and to evaluate whether additional options might be warranted. This exploratory step aimed to determine whether participants’ reported experiences were missed in the current item sets.

All preprocessing and analyses were conducted in SAS version 9.4 and R version 4.5.1.

## Results

### Sample Characteristics

Of the 335 participants enrolled at baseline, 240 participated in the EMA study.^1^ EMA participants did not differ from nonparticipants on demographic characteristics. Overall EMA compliance was 72% (*SD* = 24%, range = 0–100%). Of the 240 EMA participants, 19 were excluded for not completing the GCPS-R, and an additional 30 for responding to fewer than 50% of EMA prompts, yielding 188 participants with valid baseline EMA data. Based on their GCPS-R scores, participants were classified as having no (*n* = 29), mild (*n* = 52), bothersome (*n* = 41), or HICP (*n* = 66). The analytic sample consisted of the latter two groups (*n* = 107).

Demographic characteristics of the bothersome chronic pain and HICP groups are presented in Table 1. The HICP group was significantly younger than the bothersome chronic pain group, *t*(103.24) = 2.76, *p* = .007, but did not differ by sex, ethnicity, race, or education level (all *p*s > .18). For completeness, descriptive characteristics across all GCPS-R categories are presented in Supplementary Table 1.

**Table 1.** Demographic characteristics by Graded Chronic Pain Scale-Revised grade.

| Characteristic | Bothersome CP | HICP | *p* |
| --- | --- | --- | --- |
| Sample size, *n* | 41 | 66 |  |
| Age, mean (*SD*) | 59.59 (11.20) | 52.33 (15.89) | .007 |
| Female, % | 78.0 | 69.7 | .471 |
| Hispanic, % | 7.3 | 13.6 | .489 |
| Race, % |  |  | .367 |
| White | 61.0 | 73.8 |  |
| Asian | 26.8 | 16.9 |  |
| Other | 12.2 | 9.2 |  |
| Education, % |  |  | .874 |
| High school or less | 5.0 | 7.6 |  |
| Some college and college graduate | 65.0 | 63.6 |  |
| Advanced degree | 30.0 | 28.8 |  |
| Married/living together, % | 60.6 | 43.9 | .189 |
| Income, % |  |  | .607 |
| <\$50k | 25.6 | 32.8 | |
| \$50–99k | 25.6 | 28.1 | |
| ≥\$100k | 48.7 | 39.1 | |

*Note.* CP = chronic pain; HICP = high-impact chronic pain; *SD* = standard deviation.
*p*-values from *t*-test (continuous) or chi-square (categorical).

### Group Comparisons

#### Single Continuous Scores

Descriptive statistics and group comparisons for compliance and continuous outcome measures are presented in Table 2. Compliance with momentary and morning prompts did not differ between the groups, and average completion rates exceeded 80%.

**Table 2.**
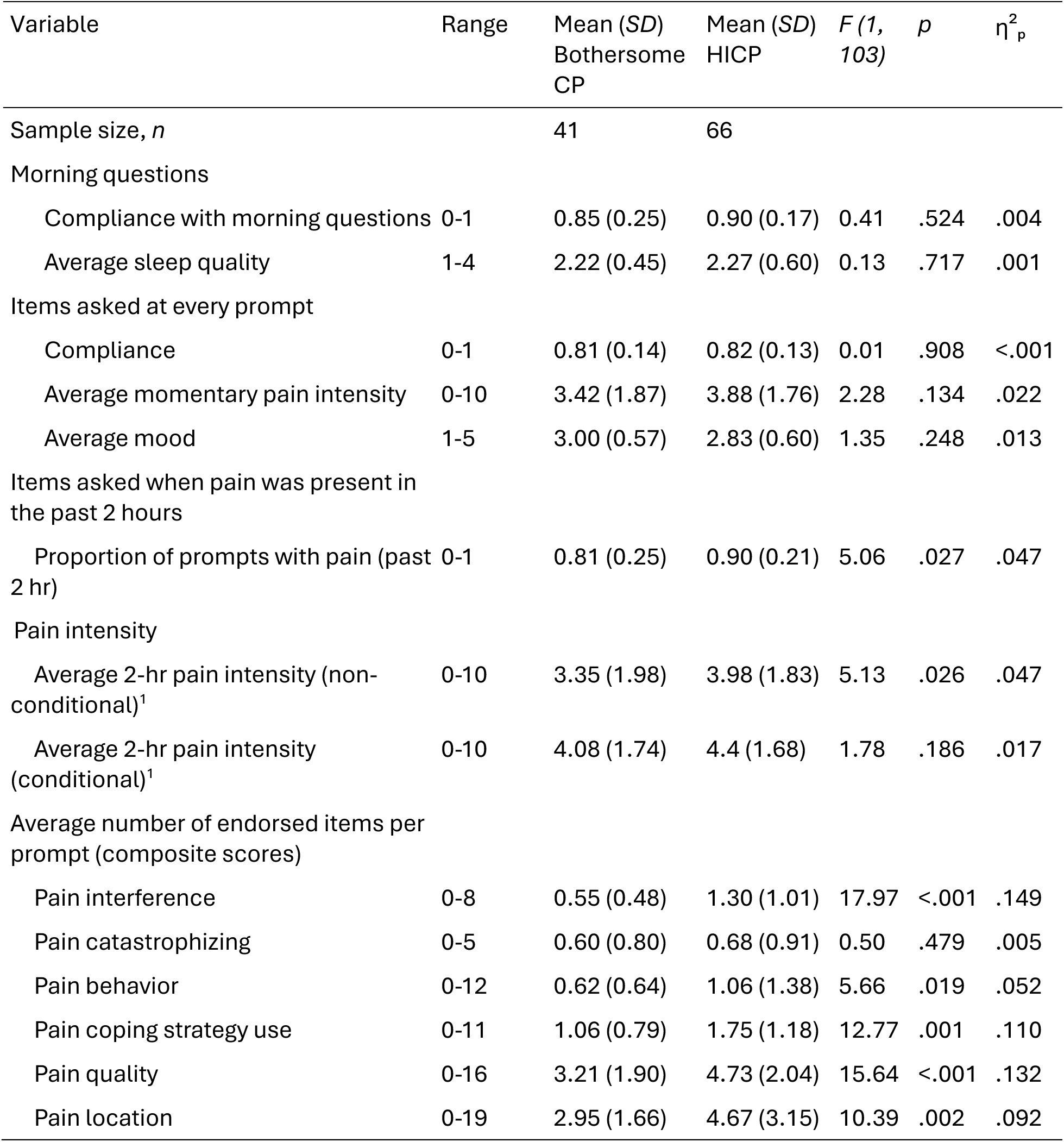
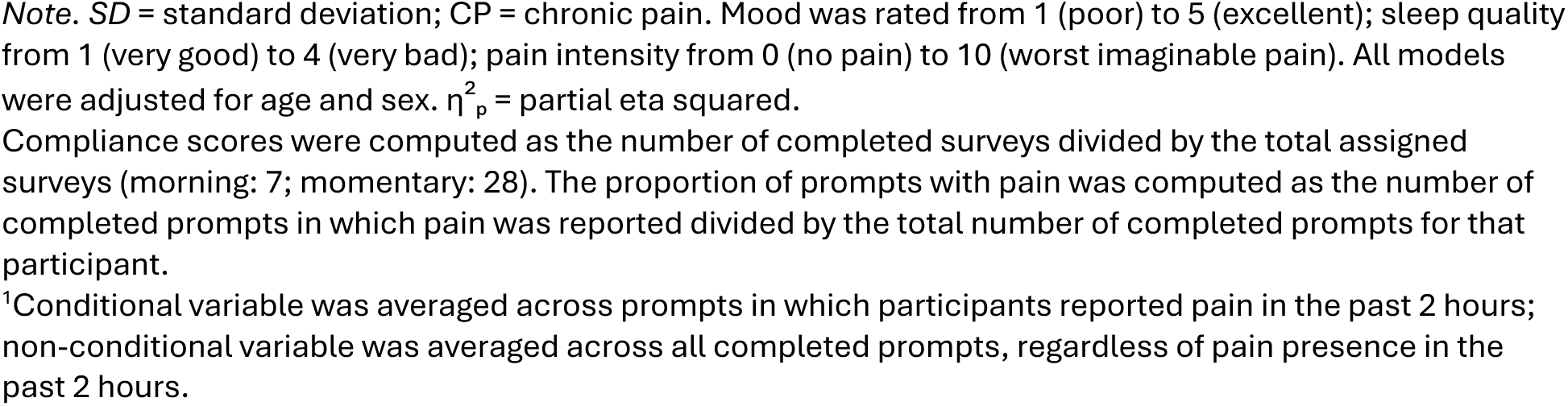
Descriptive statistics (means and standard deviations) and group comparisons for compliance and continuous outcome measures derived from the comprehensive Ecological Momentary Assessment of pain (cEMAp) in the bothersome chronic pain and high-impact chronic pain (HICP) groups.

| Variable | Range | Mean (SD)<br>Bothersome<br>CP | Mean (SD)<br>HICP | *F* (1,<br>103) | *p* | $\eta^2_p$ |
| --- | --- | --- | --- | --- | --- | --- |
| Sample size, *n* |  | 41 | 66 |  |  |  |
| Morning questions |  |  |  |  |  |  |
| Compliance with morning questions | 0-1 | 0.85 (0.25) | 0.90 (0.17) | 0.41 | .524 | .004 |
| Average sleep quality | 1-4 | 2.22 (0.45) | 2.27 (0.60) | 0.13 | .717 | .001 |
| Items asked at every prompt |  |  |  |  |  |  |
| Compliance | 0-1 | 0.81 (0.14) | 0.82 (0.13) | 0.01 | .908 | <.001 |
| Average momentary pain intensity | 0-10 | 3.42 (1.87) | 3.88 (1.76) | 2.28 | .134 | .022 |
| Average mood | 1-5 | 3.00 (0.57) | 2.83 (0.60) | 1.35 | .248 | .013 |
| Items asked when pain was present in the past 2 hours |  |  |  |  |  |  |
| Proportion of prompts with pain (past 2 hr) | 0-1 | 0.81 (0.25) | 0.90 (0.21) | 5.06 | .027 | .047 |
| Pain intensity |  |  |  |  |  |  |
| Average 2-hr pain intensity (non-conditional)<sup>1</sup> | 0-10 | 3.35 (1.98) | 3.98 (1.83) | 5.13 | .026 | .047 |
| Average 2-hr pain intensity (conditional)<sup>1</sup> | 0-10 | 4.08 (1.74) | 4.4 (1.68) | 1.78 | .186 | .017 |
| Average number of endorsed items per prompt (composite scores) |  |  |  |  |  |  |
| Pain interference | 0-8 | 0.55 (0.48) | 1.30 (1.01) | 17.97 | <.001 | .149 |
| Pain catastrophizing | 0-5 | 0.60 (0.80) | 0.68 (0.91) | 0.50 | .479 | .005 |
| Pain behavior | 0-12 | 0.62 (0.64) | 1.06 (1.38) | 5.66 | .019 | .052 |
| Pain coping strategy use | 0-11 | 1.06 (0.79) | 1.75 (1.18) | 12.77 | .001 | .110 |
| Pain quality | 0-16 | 3.21 (1.90) | 4.73 (2.04) | 15.64 | <.001 | .132 |
| Pain location | 0-19 | 2.95 (1.66) | 4.67 (3.15) | 10.39 | .002 | .092 |

*Note.* *SD* = standard deviation; CP = chronic pain. Mood was rated from 1 (poor) to 5 (excellent); sleep quality from 1 (very good) to 4 (very bad); pain intensity from 0 (no pain) to 10 (worst imaginable pain). All models were adjusted for age and sex. $\eta^2_p$ = partial eta squared.
Compliance scores were computed as the number of completed surveys divided by the total assigned surveys (morning: 7; momentary: 28). The proportion of prompts with pain was computed as the number of completed prompts in which pain was reported divided by the total number of completed prompts for that participant.
<sup>1</sup>Conditional variable was averaged across prompts in which participants reported pain in the past 2 hours; non-conditional variable was averaged across all completed prompts, regardless of pain presence in the past 2 hours.

##### Pain Intensity

The cEMAp assessed pain intensity with two timeframes: momentary (“right now”) and, when any pain was reported during the past 2 hours, over the past 2 hours (Table 2). Average momentary pain intensity across the week did not differ significantly between groups, *F*(1, 103) = 2.28, *p* = .13, η²ₚ = .02.

Individuals in the HICP group reported experiencing pain in the past 2 hours more frequently across the week than those in the bothersome chronic pain group, *F*(1, 103) = 5.06, *p* = .027, η²ₚ = .05. When restricting analyses to prompts in which pain was present (conditional), weekly average pain intensity did not differ between the two groups, *F*(1, 103) = 1.78, *p* = .19, η²ₚ = .02. However, when averaging across all completed prompts (thus incorporating both pain frequency and intensity; non-conditional), individuals in the HICP group reported higher weekly average pain intensity (*M* = 3.98, *SD* = 1.83) compared to those in the bothersome chronic pain group (*M* = 3.35, *SD* = 1.98), *F*(1, 103) = 5.13, *p* = .026, η²ₚ = .05.

##### Mood

Mood ratings were collected at every prompt and referred to the person’s mood during the past 2 hours. Average mood ratings across the week did not differ significantly between the two groups (see Table 2).

##### Sleep

Sleep quality was assessed once daily at the first completed prompt. Average sleep quality across the week did not differ significantly between the two groups (see Table 2).

#### Composite Scores

People in the HICP group reported a higher average number of endorsed pain interference, behavior, coping, quality, and location items per prompt compared to those in the bothersome chronic pain group, all *p*s < .05 (Table 2). No group differences were observed for pain catastrophizing items.

#### Multivariate Analyses

While composite scores provided a summary of overall domain endorsement, the following analyses examined whether specific pain-related experiences within each domain differed between groups. To test for overall multivariate group effects at the item level, a series of MANCOVAs were conducted for each domain, with the set of item-level proportions entered simultaneously as dependent variables and age and sex included as covariates. For domains showing a significant overall multivariate effect, follow-up ANCOVAs examined group differences in each individual item, with *p*-values adjusted for multiple comparisons. Table 3 presents the omnibus multivariate results for each domain, and follow-up ANCOVA results are visualized in Figure 1. Results from permutation- and parametric-based analyses were largely consistent, with one exception: for the pain location domain, the permutation-based test reached significance whereas the parametric MANCOVA did not.

**Figure 1.**
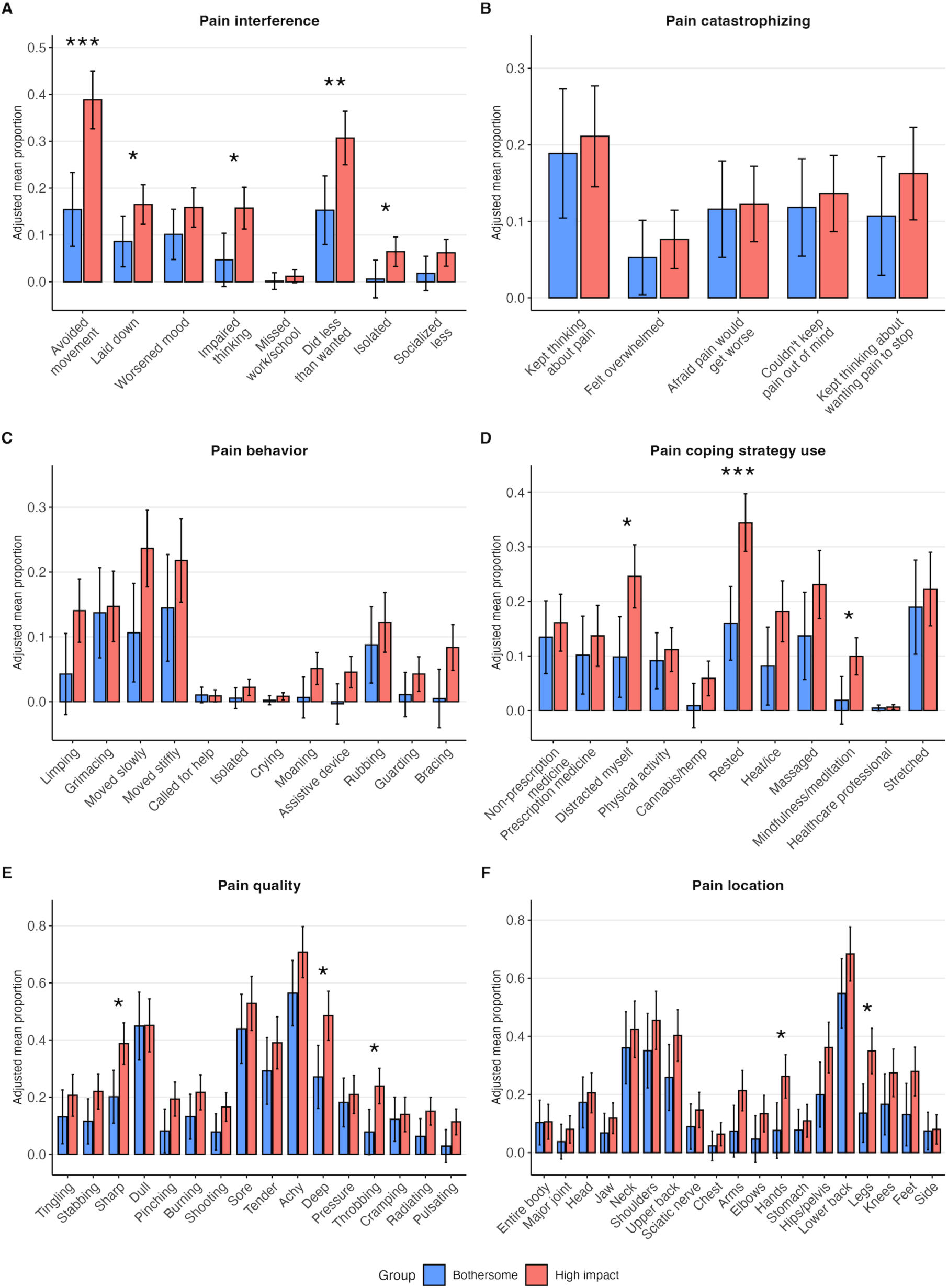
cEMAp pain domains by group. Adjusted weekly mean proportions of endorsed items for pain interference (A), pain catastrophizing (B), pain behavior (C), pain coping strategy use (D), pain quality (E), and pain location (F) in the bothersome (blue) and high-impact (red) chronic pain groups. Group differences were adjusted for age and sex. Error bars represent 95% confidence intervals. Asterisks indicate significant FDR-adjusted group differences (\**p* < .05, \*\**p* < .01, \*\*\**p* < .001).

**Table 3.** Multivariate (MANCOVA/PERMANOVA) results by the comprehensive Ecological Momentary Assessment of pain (cEMAp) domain comparing the bothersome chronic pain and high-impact chronic pain (HICP) groups.

| cEMAp domain | Number of items | Pillai's Trace | *F* | *df1* | *df2* | *p* | PERMANOVA *p* |
| --- | --- | --- | --- | --- | --- | --- | --- |
| Pain interference | 8 | .218 | 3.35 | 8 | 96 | .002 | <.001 |
| Pain catastrophizing | 5 | .018 | 0.36 | 5 | 99 | .878 | .669 |
| Pain behavior | 12 | .223 | 2.20 | 12 | 92 | .018 | .022 |
| Pain coping strategy use | 11 | .238 | 2.63 | 11 | 93 | .006 | <.001 |
| Pain quality | 16 | .275 | 2.08 | 16 | 88 | .016 | .002 |
| Pain location | 19 | .210 | 1.19 | 19 | 85 | .284 | .004 |

*Note.* *df* = degrees of freedom. All models were adjusted for age and sex. *p*-values refer to omnibus multivariate effects of group (bothersome chronic pain vs. HICP); PERMANOVA *p*-values are based on 9,999 permutations.

##### Pain Interference

The bothersome chronic pain and HICP groups differed in their patterns of pain interference, as indicated by the significant multivariate omnibus test (Table 3). Follow-up ANCOVAs revealed that, compared to the bothersome chronic pain group, individuals in the HICP group reported higher weekly proportions of prompts involving *avoiding movement/being more sedentary*, *F*(1, 103) = 20.82, *p*_adj_ < .001, η²ₚ = .17, *lying down/going to bed*, *F*(1, 103) = 5.04, *p*_adj_ = .045, η²ₚ = .05, *impaired thinking/concentration*, *F*(1, 103) = 8.90, *p*_adj_ = .011, η²ₚ = .08, *doing less than wanted*, *F*(1, 103) = 10.51, *p*_adj_ = .008, η²ₚ = .09, and *isolating/cancelling plans*, *F*(1, 103) = 4.95, *p*_adj_ = .045, η²ₚ = .05. These effects are visualized in Figure 1A, which displays adjusted weekly mean proportions (±95% CI) for each interference item by group.

##### Pain Catastrophizing

The multivariate omnibus test for pain catastrophizing was not significant, indicating no overall differences across pain catastrophizing items between the bothersome chronic pain and the HICP groups (Table 3). Accordingly, follow-up ANCOVAs were not performed. For descriptive purposes, Figure 1B displays adjusted weekly mean proportions to illustrate the frequency of each response across groups.

##### Pain Behavior

The multivariate omnibus test for pain behavior was significant, indicating overall differences between the bothersome chronic pain and HICP groups across pain behavior items over the week (Table 3). However, none of the follow-up ANCOVAs remained statistically significant after adjusting for multiple comparisons (Figure 1C). Unadjusted effects suggested that people in the HICP group more frequently reported *limping*, *F*(1, 103) = 5.78, *p* = .018, *p*_adj_ = .054, η²ₚ = .05; *moving slowly*, *F*(1, 103) = 6.92, *p* = .010, *p*_adj_ = .054, η²ₚ = .06; *using an assistive device*, *F*(1, 103) = 5.91, *p* = .017, *p*_adj_ = .054, η²ₚ = .05; and *bracing*, *F*(1, 103) = 7.20, *p* = .008, *p*_adj_ = .054, η²ₚ = .07.

##### Pain Coping Strategy Use

Multivariate analysis revealed significant group differences in pain coping strategy use over the sampling week (Table 3). Follow-up ANCOVAs showed that, compared to individuals in the bothersome chronic pain group, those in the HICP group more frequently reported *distracting themselves*, *F*(1, 103) = 9.45, *p*_adj_ = .016, η²ₚ = .08; *lying down, resting, or relaxing*, *F*(1, 103) = 17.62, *p*_adj_ < .001, η²ₚ = .15; and *using mindfulness or meditation*, *F*(1, 103) = 8.13, *p*_adj_ = .018, η²ₚ = .07. These effects are visualized in Figure 1D.

##### Pain Quality

Pain quality descriptors differed significantly between the bothersome chronic pain and HICP groups, as indicated by the multivariate omnibus test (Table 3). Follow-up ANCOVAs revealed that individuals in the HICP group more frequently described pain as *sharp*, *F*(1, 103) = 9.53, *p*_adj_ = .016, η²ₚ = .08; *deep*, *F*(1, 103) = 8.94, *p*_adj_ = .016, η²ₚ = .08; and *throbbing*, *F*(1, 103) = 9.74, *p*_adj_ = .016, η²ₚ = .09. Figure 1E presents adjusted weekly mean proportions for each pain quality item across groups.

##### Pain Location

While the parametric multivariate test for pain location did not reach significance (*p* = .284), the permutation-based test demonstrated significant group differences across body locations (Table 3). Follow-up ANCOVAs revealed consistent results between parametric and permutation-based tests for individual items. After FDR correction, the HICP group more frequently reported pain in *legs/thighs/glutes*, *F*(1, 103) = 10.78, *p*_adj_ = .019, η²ₚ = .09, and in *hands/wrists/fingers*, *F*(1, 103) = 8.96, *p*_adj_ = .028, η²ₚ = .08. Adjusted weekly mean proportions for each pain location are shown in Figure 1F.

#### Analysis of Distributional Overlaps

To complement mean-level group comparisons, we examined the degree of distributional overlap between the bothersome chronic pain and HICP groups. The overlap index quantifies the proportion of shared area between two distributions, with lower values indicating greater separation and higher values indicating greater similarity between groups. Density plots with corresponding overlap indices are presented in Figure 2A for the single continuous measures (pain intensity ratings, sleep quality, mood) and in Figure 2B for composite measures of cEMAp.

**Figure 2.**
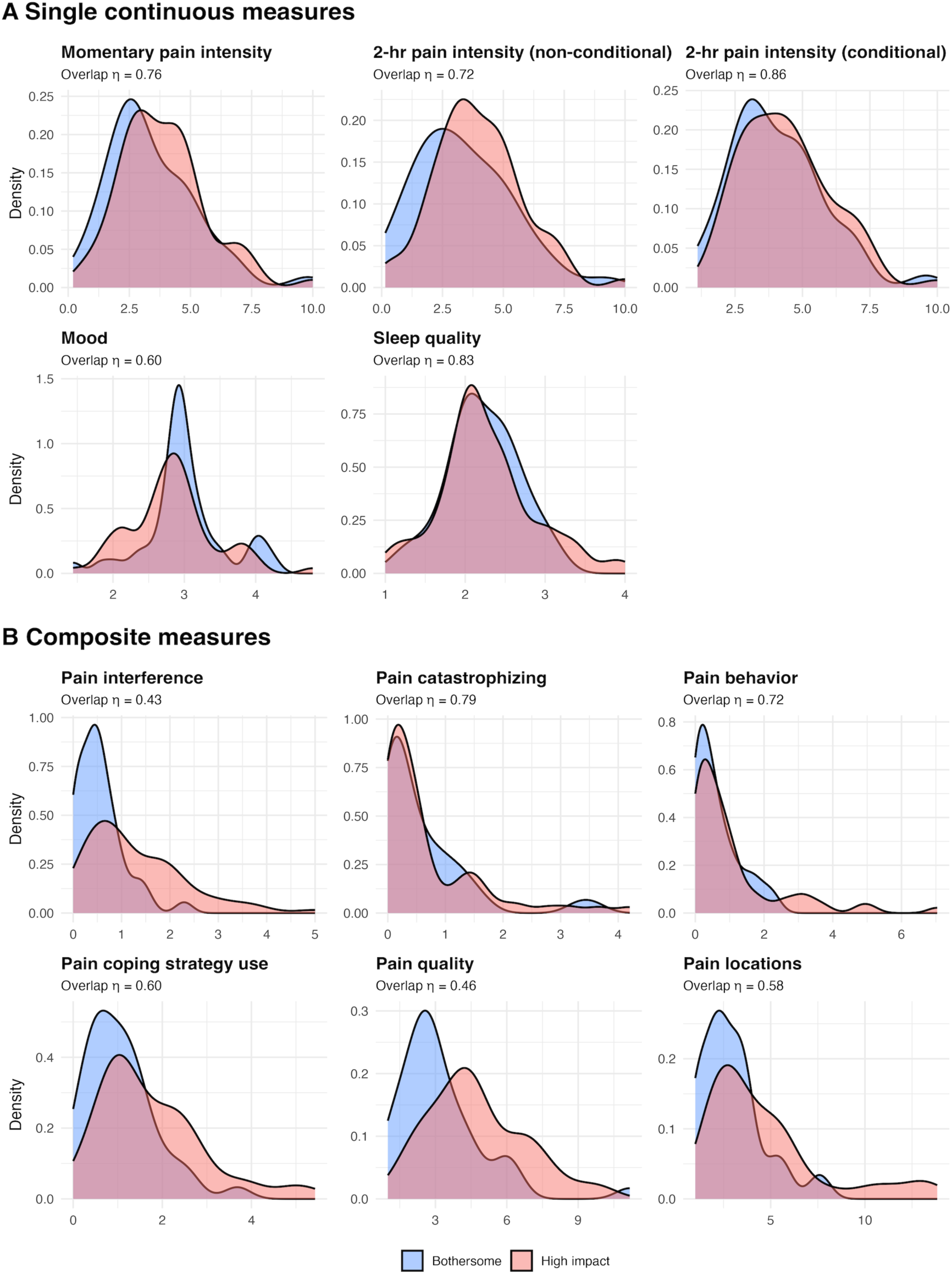
Distribution of single continuous measures (Panel A) and composite measures (Panel B) of cEMAp in the bothersome (blue) and high-impact (red) chronic pain groups. Density curves represent smoothed distributions of participant responses; overlapping regions indicate similarity between groups. The overlap coefficient (η) quantifies the proportion of shared area under the two density curves for each measure.

The overlap was the highest for pain intensity ratings (.72 - .86) and sleep quality (.83), followed by mood (.60). Across the composite cEMAp measures (Figure 2B), the mean overlap was .60, with the lowest overlap observed for pain interference (.43) and pain quality (.46) and the highest overlap for pain catastrophizing (.79).

##### Content Analysis of “Other” Responses

After adjusting for age, sex, and multiple comparisons, significant differences in the use of the “Other” response option emerged for pain interference and emotional impact. Compared to the bothersome chronic pain group, individuals in the HICP group reported higher proportions of “Other–Interference” (.08 vs .02), *F*(1, 103) = 7.97, *p*_adj_ = .024, η²p = .07, and “Other–Emotional Impact” responses (.15 vs .05), *F*(1, 103) = 7.25, *p*_adj_ = .024, η²p = .07. No group differences were observed for the remaining domains.

For pain interference, 133 “Other” responses were reviewed. Themes not captured by the existing response options included moving or working more slowly or cautiously, sleep disruption, difficulty completing daily activities and chores, and adjusting body position to avoid pain. Less common themes included resting or taking breaks, engaging in pain relief activities, and needing to stand up, stretch, or move around.

For emotional impact, 202 unique “Other” responses were coded. Approximately one third reflected negative effects on mood or well-being. Many responses also described negative thoughts or feelings about pain-related limitations, cognitive burden (e.g., distraction, difficulty concentrating, increased pain awareness), and thinking about how to lessen or relieve pain.

## Discussion

Current understanding of HICP largely stems from cross-sectional studies that collected retrospective pain data. Such studies offer limited insight into how pain and pain-related constructs manifest in daily life. To increase understanding of daily pain experiences, this study used cEMAp [12] to repeatedly assess 2-hour daily pain experiences of individuals classified as having bothersome chronic pain and HICP. Consistent with prior research based on retrospective measures, the HICP group showed greater pain interference in daily life than the bothersome chronic pain group, whereas pain intensity did not differ between the groups [30,32]. Contrary to previous work [30,32], no group differences were found for mood or pain catastrophizing in daily life. Exploratory analyses of other pain-related domains identified additional group differences. We also observed distributional overlap between the two groups in several domains; thus, even when mean-level group differences were present, individuals in the two groups showed some similarity in daily experiences across several domains. We next discuss these findings in more detail.

The feature most clearly distinguishing HICP from bothersome chronic pain in daily life was the frequency of interference with physical, mental, and social activities. Individuals in the HICP group reported more limitations per prompt and more frequently reported avoiding movement, spending more time being sedentary, experiencing cognitive difficulties, doing less than they wanted, and isolating socially across the sampling week. Effect sizes for these differences ranged from moderate to large, with the largest effects observed for avoiding movement and doing less than they wanted. Notably, individuals classified as having HICP avoided movement during roughly 40% of pain-related prompts and reported doing less than they wanted during about 30%. This pattern of results provides support for known-groups validity of the GCPS-R classification in daily life, as the clearest group differences emerged in the frequency of pain-related interference, the construct used in the operational definition of HICP [32]. In addition, these findings are consistent with previous retrospective studies that showed that individuals classified as having HICP more often reported life activity restrictions such as reduced physical activity and activities of daily living, limited social activities, as well as spending much of the day in bed [30,32,36].

Also consistent with prior research, average momentary pain intensity did not differ between groups, suggesting that intensity alone does not differentiate HICP from bothersome chronic pain in daily life. This was further supported by the conditional analysis of 2-hour pain intensity, which averaged intensity ratings only across prompts in which pain was present and similarly found no group differences. In contrast, the non-conditional average, which incorporates both pain intensity and pain frequency across all prompts, was significantly higher in the HICP group. This pattern suggests that the mean difference reflects more frequent pain episodes rather than greater pain intensity when pain occurs. The ability to disentangle pain frequency from pain intensity represents an important advantage of EMA over traditional retrospective measures.

In contrast, some pain-related constructs that have demonstrated differences between HICP and bothersome chronic pain in cross-sectional studies, such as poorer emotional well-being and greater pain catastrophizing [30,32], did not differ between groups when assessed in daily life using short reporting periods. These findings suggest that certain group differences previously observed in retrospective studies may be less pronounced in daily life assessments. Alternatively, differences in methodology, particularly the use of shorter versus longer reporting intervals, could have contributed to these discrepancies in findings [5,34], as prior work has shown that mean-level group differences may be larger for retrospective ratings than for momentary assessments [11]. At the same time, findings regarding greater anxiety/depression and pain catastrophizing in the HICP group have been inconsistent across studies and associated effect sizes were modest [30,32]. Future studies using larger samples are needed to further evaluate these distinctions.

Exploratory analyses across additional cEMAp domains yielded a detailed picture of how HICP manifests in daily life, extending our understanding of HICP into areas that have not been previously investigated. On average per prompt, individuals classified as having HICP reported more pain behaviors, endorsed using a greater number of coping strategies, described their pain using a wider range of quality descriptors, and indicated more pain locations than those in the bothersome chronic pain group. To further clarify these composite-level differences, we examined the frequency of individual items within each domain. Several specific items showed significant group differences. Compared to the bothersome chronic pain group, individuals in the HICP group more often reported using particular coping strategies (such as distraction, resting, or mindfulness practices) and described their pain using specific qualities (particularly sharp, deep, or throbbing sensations). However, no consistent item-level differences were observed for other pain-related experiences in daily life assessed using cEMAp. Together, these findings suggest that although HICP is associated with some distinct manifestations of pain in daily life, many day-to-day experiences are shared between the two groups.

To complement mean-level analyses and better characterize the extent of separation between groups, we examined the degree to which the distributions of pain intensity and pain-related experiences overlapped. Consistent with the observed effect sizes, overlap was lowest for domains showing the largest group differences, particularly pain interference. However, even for this domain, the distributions were not completely separated. For domains with smaller or nonsignificant mean differences, the overlap was more pronounced. Together, these findings highlight heterogeneity within each group. Of particular relevance to the core features of HICP, the findings suggest that some individuals classified as having bothersome chronic pain may experience pain-related burden in daily life comparable to those classified as having HICP.

Taken together, the everyday lives of individuals classified as having bothersome chronic pain or HICP differ most clearly in the frequency of functional interference, while showing similarities across several other domains. If replicated, these findings may have research and clinical implications. In clinical contexts, brief real-world sampling of pain-related experiences could complement retrospective classifications by providing additional information about pain-related burden and serving as an outcome measure for monitoring change. For research, future refinements of the HICP classification might benefit from integrating intensive daily sampling of pain-related experiences (e.g., for one week) as a source of information for defining subgroups of individuals with chronic pain. Relying solely on retrospective assessments at a single time point might obscure important similarities and differences between groups of patients as they go about their daily lives. Incorporating daily sampling of patients’ experiences could provide a richer and more nuanced classification scheme.

This study has several strengths. First, it is the first to examine pain-related experiences of individuals classified as having HICP and bothersome chronic pain in daily life. The use of EMA enabled a more granular, ecologically valid assessment of pain-related experiences and reduced recall bias. In addition, employing cEMAp allowed us to assess a broader set of pain-related domains than has been examined in prior work, providing a more comprehensive characterization of daily life experiences. This approach also provides a framework for future studies to investigate which momentary factors might give rise to specific pain-related limitations observed in the HICP group, such as movement avoidance or reduced activity. All analyses controlled for sex and age, factors known to influence pain reporting [13,14], and both parametric and permutation-based tests yielded largely convergent results, increasing confidence in the robustness of the findings.

Several limitations should also be noted. The sample size was modest, and the sample consisted predominantly of individuals with chronic low back pain, which limits generalizability to other pain conditions. The EMA data used for this study considered only a single week and, therefore, captures a limited snapshot of individuals’ daily experiences. Longer EMA periods or repeated sampling bursts could provide more stable estimates of daily life experiences of pain. Additionally, the “other” option for the impact on feeling/thinking item was endorsed frequently (20% of prompts), and content analysis suggested several themes that could inform refinement of future cEMAp versions. Finally, affect was assessed using a single-item measure, which may not fully capture the complexity of emotional experiences.

In summary, using one week of EMA in adults classified as having bothersome chronic pain and HICP, the most consistent day-to-day distinction between groups was the frequency of functional impact (i.e., activity limitation and interference). This aligns with the current operationalization of HICP, which primarily reflects how often pain disrupts daily functioning rather than uniformly different pain intensity or affective experience. In contrast, many other pain-related experiences showed only small differences. At the same time, examination of distributional overlap suggested some heterogeneity within groups.

This suggests that a subset of individuals classified as having bothersome chronic pain may experience HICP-like burden at least some of the time. Clinically, brief, real-world sampling of functional impact may complement retrospective measures by identifying patients whose day-to-day impairment is under-recognized by categorical screening. For research, integrating EMA-derived functional-impact phenotypes with longitudinal outcomes and biomarker data may enable more precise stratification and improve prediction of prognosis and treatment response.

## Generative AI Statement

During the preparation of this work, the first author used ChatGPT 5.2 tool to assist with language editing and clarity. All content was reviewed and verified by the authors.

## Data Availability

All data and analytical code are available upon request to the corresponding author.

## Acknowledgments

The authors would like to thank all participants for taking part in this study. This work was supported by the National Institutes of Health under NIA R37AG057685 (A.A. Stone, P.I.) and NINDS R61NS118651 (S.C. Mackey, P.I.).

Data and materials availability: All data and analytical code are available upon request to the corresponding author.

## Footnotes

1 Data collection for the main project began in April 2022. The EMA study module was added in March 2023; 47 participants enrolled before its inclusion did not complete EMA.

