## Supplementary Table 1 for "Ecological Momentary Assessments of daily pain experiences in bothersome and high-impact chronic pain"

**Supplementary Table 1.** Demographic characteristics by all grades of Graded Chronic Pain Scale-Revised.

| Characteristic | No CP | Mild CP | Bothersome CP | HICP |
|---|---|---|---|---|
| Sample size, *n* | 29 | 52 | 41 | 66 |
| Age, mean (*SD*) | 54.17 (15.54) | 60.12 (11.66) | 59.59 (11.20) | 52.33 (15.89) |
| Female, % | 75.9 | 67.3 | 78.0 | 69.7 |
| Hispanic, % | 13.8 | 13.7 | 7.3 | 13.6 |
| Race, % | | | | |
| White | 48.3 | 70.0 | 61.0 | 73.8 |
| Asian | 31.0 | 22.0 | 26.8 | 16.9 |
| Other | 20.7 | 8.0 | 12.2 | 9.2 |
| Education, % | | | | |
| High school or less | 3.4 | 0.0 | 5.0 | 7.6 |
| Some college and college graduate | 62.1 | 64.0 | 65.0 | 63.6 |
| Advanced degree | 34.5 | 36.0 | 30.0 | 28.8 |
| Married/living together, % | 51.9 | 70.5 | 60.6 | 43.9 |
| Income, % | | | | |
| <$50k | 22.2 | 6.4 | 25.6 | 32.8 |
| $50–99k | 29.6 | 36.2 | 25.6 | 28.1 |
| ≥$100k | 48.1 | 57.4 | 48.7 | 39.1 |

*Note.* CP = chronic pain; HICP = high-impact chronic pain; *SD* = standard deviation.
